# Exploring the secretion of immunogenic gluten peptides in breast milk from celiac and non celiac mothers

**DOI:** 10.1101/2024.05.22.24307399

**Authors:** Ángela Ruiz-Carnicer, Verónica Segura, María de Lourdes Moreno, Cristóbal Coronel-Rodríguez, Carolina Sousa, Isabel Comino

## Abstract

**Background:** Exposure to antigens is crucial for child immune system development, aiding disease prevention and promoting infant health. Some common food antigen proteins are found in human breast milk. However, it is unclear whether gluten antigens linked to celiac disease (CD) are transmitted through breast milk, potentially impacting the development of the infant’s immune system.

**Objective:** This study aimed to analyze the passage of gluten immunogenic peptides (GIP) into human breast milk. We evaluated the dynamics of GIP secretion after lactating mothers adopted a controlled gluten-rich diet.

**Methods:** We prospectively enrolled 96 non-CD and 23 CD lactating mothers, assessing total proteins and casein in breast milk, and GIP levels in breast milk and urine. Subsequently, a longitudinal study was conducted in a subgroup of 12 non-CD lactating mothers who adopted a controlled gluten-rich diet. GIP levels in breast milk and urine samples were assayed by multiple sample collections over 96 hours.

**Results:** Analysis of a single sample revealed that 24% of non-CD lactating mothers on a regular unrestricted diet tested positive for GIP in breast milk, and 90% tested positive in urine, with significantly lower concentrations in breast milk. Nevertheless, on a controlled gluten-rich diet and the collection of multiple samples, GIP were detected in 75% and 100% of non-CD participants in breast milk and urine, respectively. The transfer dynamics in breast milk samples were long-enduring and GIP secretion persisted from 0 to 72 h. In contrast, GIP secretion in urine samples was limited to the first 24 h, with inter-individual variations. In the cohort of CD mothers, 82.6% and 87% tested negative for GIP in breast milk and urine, respectively.

**Conclusions:** This study definitively established the presence of GIP in breast milk, with substantial inter-individual variations in secretion dynamics. Our findings provide insights into distinct GIP kinetics observed in sequentially collected breast milk and urine samples, suggesting differential gluten metabolism patterns depending on the organ or system involved. Future research is essential to understand whether GIP functions as sensitizing or tolerogenic agents in the immune system of breastfed infants.

## Introduction

Celiac disease (CD) is a systemic disorders triggered by exposure to dietary gluten in genetically predisposed individuals [1,2]. It is a common disease that can affect individuals of all ages and is characterized by a wide spectrum of clinical manifestations. The symptoms include abdominal pain, bloating, nausea, vomiting, and/or diarrhea. Additionally, extra-intestinal manifestations can occur, such as blistering skin rashes, ataxia, bone disease, and issues in the reproductive and endocrine systems, among others [3].

The diagnostic rate for this pathology has increased over the last 10 years [4]. Worldwide epidemiological data show that CD is ubiquitous, with a prevalence of 1.4% [5], and is higher in females than in males [4–8]. The increase in this prevalence may, in part, be attributed to improved recognition and testing of the disease. However, there may also be a real increase in the incidence of this immune disorder, related to environmental factors that promote a loss of tolerance to dietary gluten [9]. Breastfeeding and the timing of gluten introduction have been particularly considered influential factors [10]. Nevertheless, the available evidence regarding the relationship between CD and breast milk remains inconclusive. Extended breastfeeding and, notably, breastfeeding at the time of gluten introduction seem to decrease the risk of developing CD or at least postpone its onset [11,12]. However, conflicting findings persist, as some studies have been unable to establish breastfeeding as protective against the development of CD [13–15].

Breastfeeding significantly influences the composition of the intestinal microbiota in the infant and offers substantial potential to prevent allergic diseases [16,17]. The study of breast milk composition is crucial not only for understanding the nutritional requirements of infants, but also for evaluating its impact on neurodevelopment, immune system maturation, and gut development, among others [18]. Additionally, breastfeeding plays a dual role in promoting tolerance through the presence of immunomodulatory substances and facilitating antigen transfer. The presence of food antigens in breast milk is a significant source of early oral exposure that is critical for establishing and reinforcing oral tolerance [19].

Common food antigens found in human breast milk include proteins derived from egg (ovalbumin) [20], cow milk (casein and beta-lactoglobulin) [21], and peanuts (arah h1 and arah h2) [22,23]. The frequency and concentration of these dietary proteins in breast milk vary widely among women, with approximately 50% excreting them at concentrations ranging from 0.1 to over 1000 ng/ml [24–26]. However, the presence of certain proteins/peptides including gluten remains unclear [27,28]. Gluten proteins have unique properties compared to other dietary proteins. In particular, they contain high proportions of glutamine and proline residues, making them resistant to degradation by gastrointestinal proteases. Consequently, relatively long gluten peptides with intact immunotoxic epitopes accumulate in the lumen of the small intestine and cross the epithelial barrier, ultimately entering the systemic circulation [29–31]. This raises the possibility of their subsequent secretion through the mammary glands into breast milk. Therefore, the possible presence of gluten peptides in breast milk could promote oral tolerance or sensitivity to gluten in breastfed infants.

Our research group is a pioneer in identifying and analyzing gluten in human samples. In particular, monoclonal antibodies (moAbs) have been used to develop immunoassays for the sensitive and specific detection of the gluten immunogenic peptides (GIP). These assays have enabled to measure GIP concentrations in human samples, including feces and urine, confirming their absorption into the bloodstream [32–36]. Given their resistance to gastrointestinal digestion, GIP play a key role in triggering immunogenic responses within T cells of individuals diagnosed with CD. However, to date, there have been no studies that confirm the existence of gluten peptides in breast milk and if they are immunogenic. On this basis, the aim of this study is to understand the transfer and secretion dynamics of GIP in breast milk. Initially, we carried out a cross-sectional study to analyze the GIP in breast milk samples within a cohort of mothers with and without CD and assessed total protein and caseins secretion. Secondly, we conducted an exploratory longitudinal clinical study to elucidate the differential kinetics of GIP secretion in human milk and urine by examining non-CD mothers after a controlled gluten-rich diet.

## Materials and methods

### Study design and participants

#### Cross-sectional study

This cross-sectional study was conducted between September 2020 and September 2023, and included lactating mothers with and without CD. Potential volunteers were invited to participate via the following official notification: Centro de Salud Amante Laffón (Sevilla, Spain), Federación de Asociaciones de Celíacos de España (FACE), Asociación Provincial de Celíacos de Sevilla (Asprocese) and social networks (https://celiacos.org/la-universidad-de-sevilla-estudia-la-presencia-de-peptidos-del-gluten-en-la-leche-materna/).

A questionnaire was administered to assess the gestational age of the children at delivery and during sample collection process. Furthermore, mothers with CD were asked about the date of CD diagnosis, results of the last duodenal biopsy study, and serum CD antibodies. The inclusion criteria for the cohort of non-CD mothers were as follows: (1) over 18 years of age; (2) regular gluten consumption; and (3) no prior diagnosis of CD, non-CD gluten sensitivity, food allergies, food intolerances, or other gastrointestinal diseases. The inclusion criteria for the cohort of mothers with CD were as follows: (1) over 18 years of age; (2) breastfeeding mothers previously diagnosed with CD, and (3) all volunteers had followed a gluten-free diet (GFD) for at least 2 years prior to enrolling in the study. The exclusion criteria were as follows: (1) participants with associated pathologies or severe psychiatric diseases; and (2) participants who did not collect the samples properly.

This study was conducted in accordance with the principles of the Declaration of Helsinki. The study protocol was approved by the regional research ethics committee (ID/Numbers: 0364-N-20, Comunidad Autónoma de Andalucía, Spain), and written informed consent was obtained from all participants.

Detailed instructions were provided to all participants at the beginning of the study. Breast milk and urine samples were consecutively collected from each participant. The parameters analyzed were total protein, casein and GIP level in breast milk, and GIP level in urine.

#### Longitudinal study

A longitudinal study was conducted in a selected subgroup of non-CD mothers who were instructed to complete a two-phase study: (1) an initial 3-day phase with a controlled gluten-rich diet and (2) a second 5-day phase with a strict GFD. During this study, participants were provided with a menu planning with foods rich in gluten (pizza, sandwich, wheat pancakes, pasta, croutons, lasagna, etc.) for the first period, and menus with gluten-free (unprocessed or minimally processed) foods for the second phase (Figure 1). All volunteers collected consecutive urine and milk samples at 0 h during the controlled gluten-rich diet and at 3, 6, 24, 48, 72, and 96 h after the start of the GFD.

**Figure 1.**
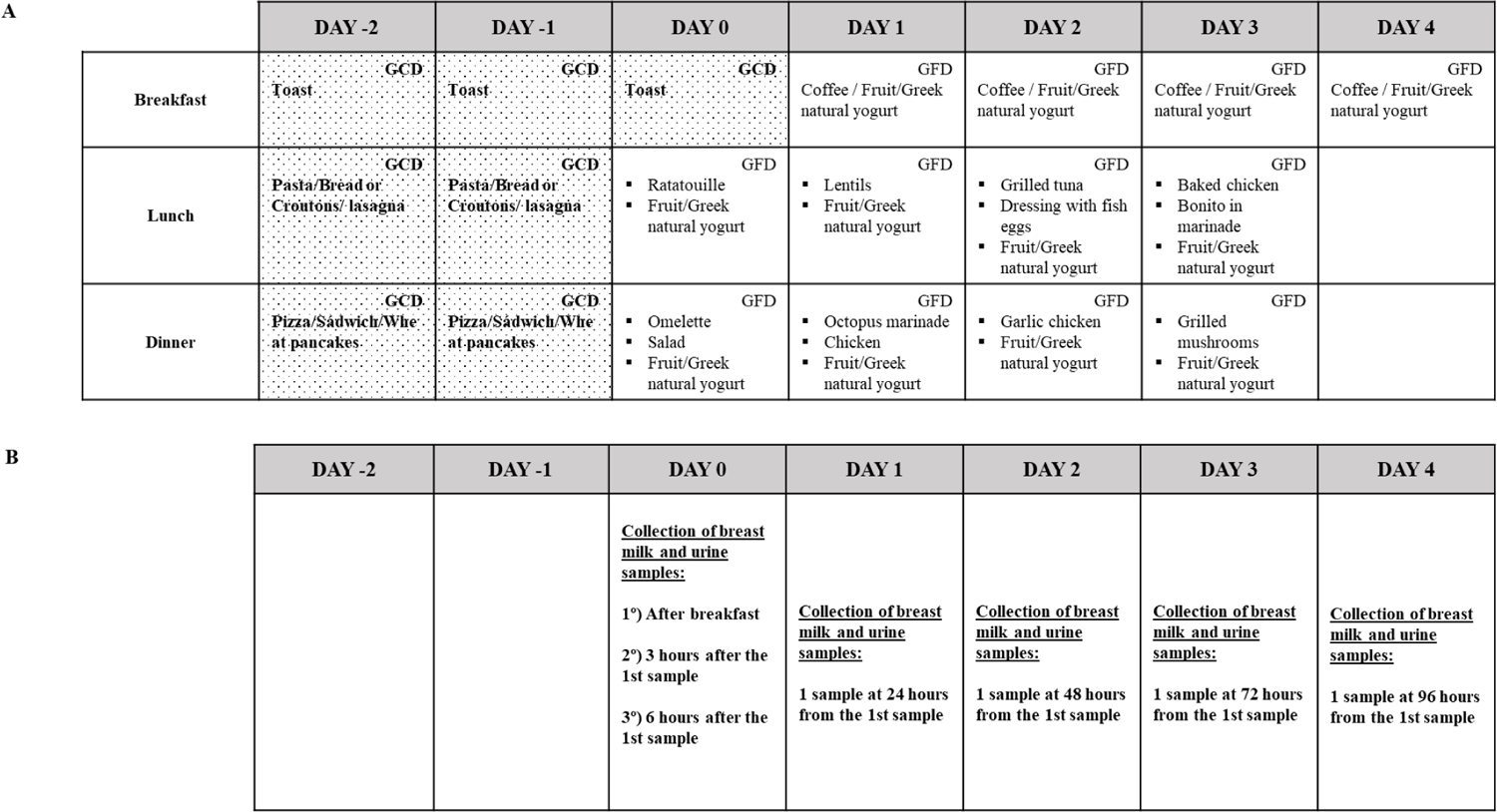
Menu planning and sample collection planning. (A) Menu planning with foods rich in gluten and menus with gluten-free. (B) Sample collection planning. GCD, gluten-containing diet; GFD, gluten-free diet.

### Milk and urine collection

The participants were instructed to collect 10 mL of mature milk and 5 mL of urine in a sealed sterile container. Mothers expressed the milk samples manually or by pump and immediately froze the milk and urine sample at −20 °C until collected by the researchers. Once in the laboratory, the samples were homogenized, and after aliquoting, they were stored at −20 °C. To optimize the milk protocol, the following were used: 1) whole breast milk samples, and 2) whey samples obtained by centrifuging an aliquot of the milk sample for 5 min at 8000 x *g* (removal of fat content and cellular elements that could interfere with immunoassays). To ensure temporal coherence and minimize potential data variations, milk and urine samples were collected consecutively over time.

### Total protein concentration

The protein concentration in the whole milk and whey was quantified using a Pierce BCA Protein Assay Kit (Thermo Fisher Scientific Inc. Waltham, Massachusetts, USA) adapted to microplates. Briefly, 25 μL of milk/whey or standard diluted was added to 96-well plates in duplicate. After covering the plates and incubating at 37°C for 30 minutes, optical densities were read at 562 nm in the Multiskan Sky Reader (Thermo Fisher Scientific Inc. Waltham, Massachusetts, USA). Each sample was participants to duplicate analyses on different dates.

### Casein concentration

The casein concentration in breast milk samples was measured by sandwich-type enzyme-linked immunosorbent assay (ELISA) using the AlerTox ELISA Casein Kits (AlerTox ELISA, Hygiena Diagnostic España S.L., Seville, Spain), following the manufacturer’s guidelines with minor modifications. Briefly, breast milk samples were incubated for 15 min at 60°C with gentle agitation in extraction buffer and centrifuged. ELISA was performed using a microtiter plate and standards previously diluted 1:100. The samples were then incubated with a casein-conjugated antibody for 20 min. The TMB casein substrate solution was then added. Colour development was stopped with a stop solution of casein, and absorbance at 450 nm was measured using a Multiskan Sky Reader microplate reader (Thermo Fisher Scientific Inc., Waltham, Massachusetts, USA). A minimum of two separate aliquots from each sample were examined on different days.

### Determination of GIP in milk and urine samples

Milk and urine samples were processed according to the guidelines provided by the manufacturer (iVYCHECK GIP Urine based on G12 and A1 moAbs; Biomedal S.L., Seville, Spain).

Subsequently, 100 µL of each sample was applied to the detection test strip. After a 30-minute incubation period, the immunochromatographic strip was assessed using a lateral flow test reader cassette. The limit of detection (LoD) was established through visual examination (2.25 ng GIP /mL), and the measuring range was 3.12-25 ng GIP /mL that was determined using the iVYCHECK Reader (Biomedal S.L.). Each sample was run in duplicate, and at least two different aliquots of each sample were tested on different days.

### Statistical Analysis

The results of the quantitative variables were expressed using the median and interquartile range (IQR), and those of the qualitative variables were expressed as percentages. The goodness-of-fit to normality was calculated using the Shapiro–Wilk test.

The Mann–Whitney U test was used to compare quantitative variables in independent groups, and the Wilcoxon signed-rank test was used to compare quantitative variables in dependent groups. The McNemar test was used to compare qualitative variables within dependent groups. For all the cases, P < 0.05 was considered statistically significant. The data analysis was performed using SPSS 26.0 software for Windows (SPSS Inc.).

## Results

### Participants

We recruited 96 non-CD lactating mothers taking a normal unrestricted diet with a median age of 35 (IQR: 32-37) and 23 CD lactating mothers under long-term GFD with a median age of 35 (IQR: 31-37). In non-CD mothers, the mode of delivery of the fetus was: 81% (78/96) vaginal and, 17% (16/96) cesarean. For CD mothers, the mode of delivery of the fetus was: 17% (4/23) vaginal and, 4% (1/23) cesarean. The gestational age, 40 (IQR: 39-40) weeks, was similar in both cohorts. However, the median of the babýs age was 2 (IQR:1-5) months in non-CD mothers and 8 (IQR: 4-20) months in CD-mothers.

### Cross-sectional study

As this was a non-interventional study, and neither the amount of fluid ingested, nor the feeding pattern of the lactating mothers was modified. Additionally, urine and breast milk samples were collected only once.

#### Determination of total protein and casein in breast milk

To evaluate whether differences in the diets of non-CD and CD mothers could affect the protein content of their breast milk, we analyzed protein concentrations before and after whey separation. The protein concentration was higher in whole milk than in whey in both cohorts (Wilcoxon test, P<0.001) (Figure 2A). In CD mothers, the median was 9.7 (IQR= 9.1-12.8) and 9 mg/mL (IQR =8-11) for whole breast milk and whey samples, respectively, while in non-CD mothers was 10.7 (IQR= 9.7-12) and 9.8 mg/mL (IQR=8.6-11.6). Protein levels values were similar between the study cohorts, with no significant differences (Mann–Whitney U test, P=0.273).

**Figure 2.**
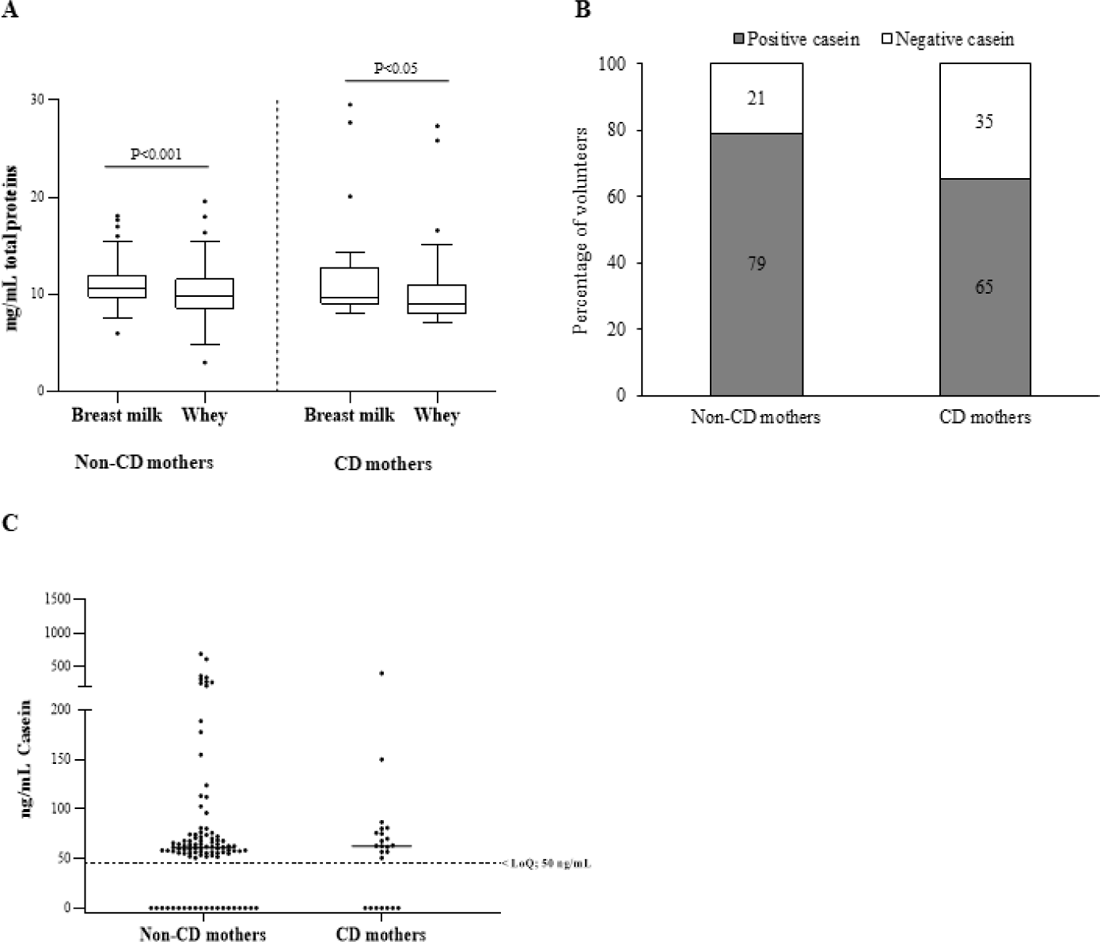
Total proteins and bovine caseins in breast milk. (A) Total protein content (mg/mL) in breast milk and whey (milk serum) of non-CD and CD mothers; (B) percentage of non-CD and CD mothers with casein in breast milk; (C) casein concentration in non-CD and CD mothers. Differences between groups were tested with a Wilcoxon signed-rank test in dependent groups and Mann–Whitney U test in independent groups. CD, celiac disease; LoQ, limit of quantification.

Bovine casein is one of the non-human proteins most frequently found in human breast milk [21,37,38]. In this study, casein was detected in 78.15% of the samples (93/119) (Figure 2B), specifically 65% (16/23) in the cohort of CD mothers and 79% (76/96) in non-CD mothers (median 62.5 ng/mL; IQR 50–76 and median 60.7 ng/mL; IQR 52.2–73.4, respectively). As expected, the results showed no significant differences between the bovine casein values of the control and CD mothers (Mann–Whitney U test, P=0.797) (Figure 2C).

#### Determination of GIP in breast milk

We investigated the presence of gluten in breast milk by quantifying GIP using specific moAbs (G12/A1). In the cohort of non-CD mothers without gluten restriction in their diet, 24% (23/96) of breast milk samples contained detectable levels of GIP. There were no significant differences in the GIP levels of whole milk and whey for these mothers. Urine samples tested positive for GIP in 90% (86 out of 96) of volunteers within this cohort. The milk and urine samples were collected consecutively over time, and our findings revealed that the concentrations of GIP in the milk were significantly lower than those in the urine (average range according to the Mann–Whitney U test: 60.71 and 132.39, respectively) (P < 0.001) (Figure 3). However, 96% (22/23) of mothers who tested positive in breast milk also had a GIP positive result in urine.

**Figure 3.**
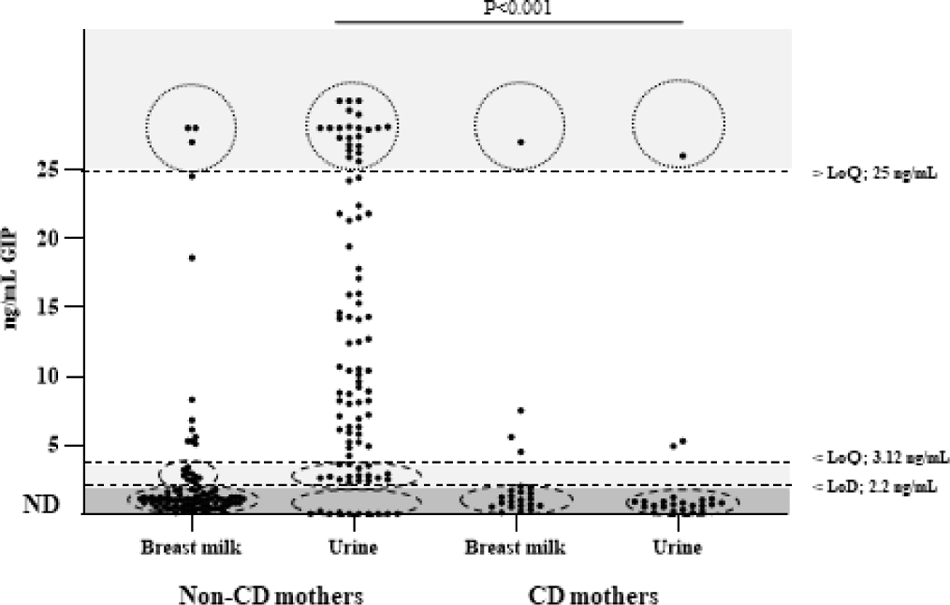
Detection and quantification of GIP in urine and breast milk of non-CD mothers and CD mothers. n = 96 (non-CD mothers), n = 23 (CD mothers). The data points include 2 values per volunteer. Differences in GIP levels in breast milk and urine between the 2 groups were tested with a Mann–Whitney U test. CD, celiac disease; GIP, gluten immunogenic peptides; LoD, limit of detection; LoQ, limit of quantification; ND, not detectable.

In the cohort of CD mothers, 82.6% (19/23) tested negative for GIP in breast milk. GIP was detected in only four breast milk samples and three urine samples from 23 participants, and one of the participants showed positive results for both samples. No significant differences were found between the GIP values of the milk and urine samples (P > 0.05).

These results suggest the presence of GIP across both biological fluids, albeit at varying levels, underscoring the importance of comprehensive assessment methods in elucidating GIP dynamics in lactating mothers with CD.

### Longitudinal study

#### Kinetics of GIP secretion in breast milk

A parallel longitudinal descriptive study was conducted using a subgroup of 12 volunteers from the cohort of non-CD mothers. This study aimed to elucidate GIP kinetics in breast milk after maternal ingestion of gluten. For this purpose, all volunteers were instructed to complete a two-phase study: phase 1 on a controlled gluten-rich diet, and phase 2 on a strict GFD. GIP levels were quantified in breast milk and urine samples taken at various time points, from 0 to 96 h after gluten ingestion and at the onset of the GFD.

The results were GIP positive in 75% (9/12) of the participants in breast milk samples and 100% (12/12) of the urine samples (Figure 4), considering at least one positive sample per participant. After adopting a gluten-rich diet (0 h), 54.5% (6/11) of the volunteers tested positive for GIP in their breast milk. At 3 and 6 h from the beginning of the GFD, GIP was detected in 33.3% (4/12) and 16.7% (2/12) of the volunteers, respectively. In addition, we found positive GIP in breast milk on the following hours: at 24, 48 and 72 h in 16.7% (2/12), 25% (3/12) and 8.3% (1/12) of the participants, respectively. The means GIP values detected in breast milk were similar between 0 and 48 h, however, significant differences were found after 72 h (P<0.05) (Figure 5A). No GIP was detected in breast milk after 72 h in the volunteers studied. However, the period of GIP excretion in urine after the start of GFD is limited to the first 24 h (100% of urine samples at 0h, 92% at 3 and 6 h, and 17% at 24 h) (Figure 5B).

**Figure 4.**
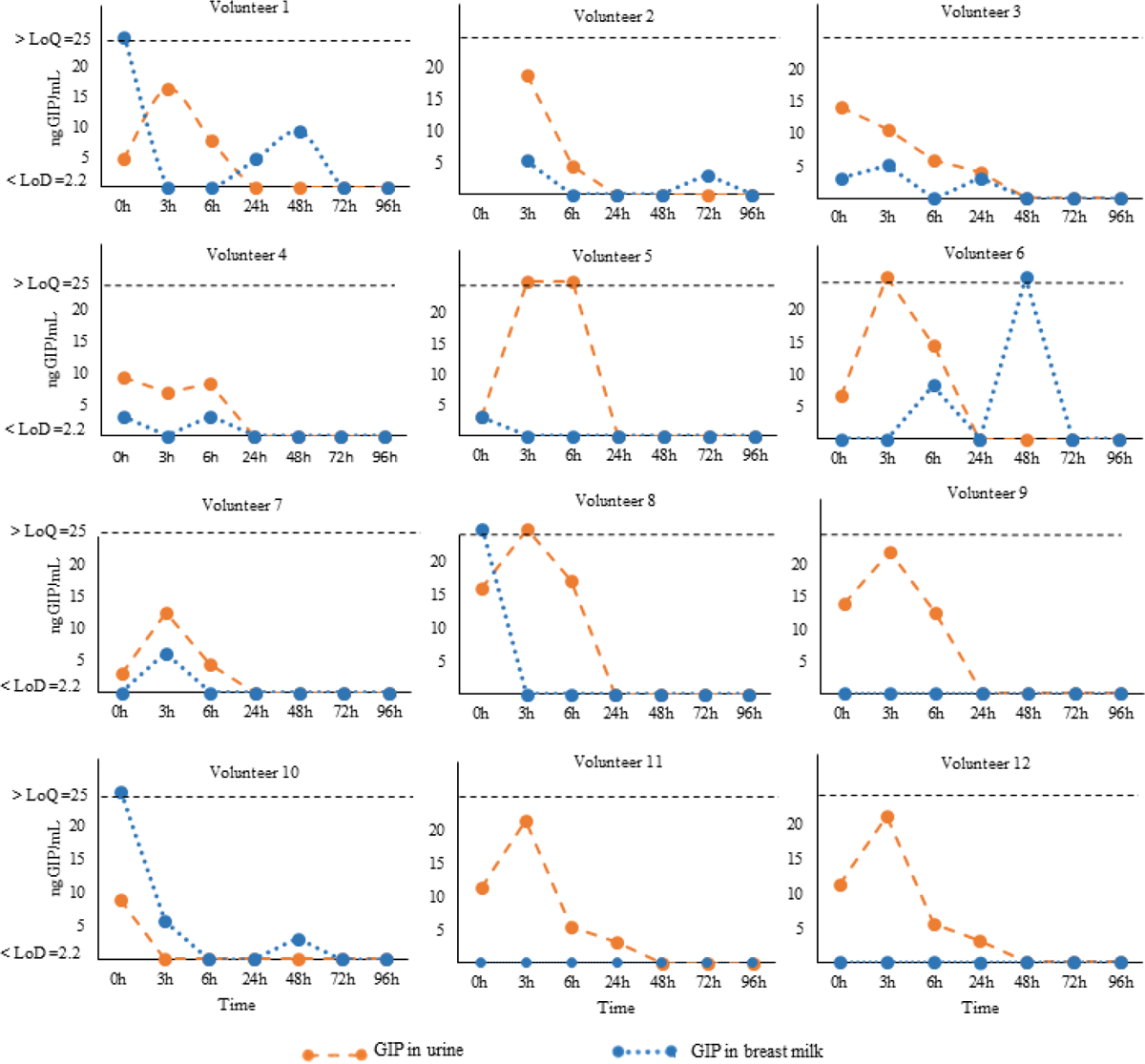
Kinetics of GIP secretion in breast milk and urine. Individual GIP excretion kinetics of 12 non-CD mothers on a GFD for over 96 h. The dashed orange lines correspond to breast milk samples, and blue lines represent urine samples. GFD, gluten-free diet; GIP, gluten immunogenic peptides; LoQ, limit of quantification; LoD, limit of detection.

**Figure 5.**
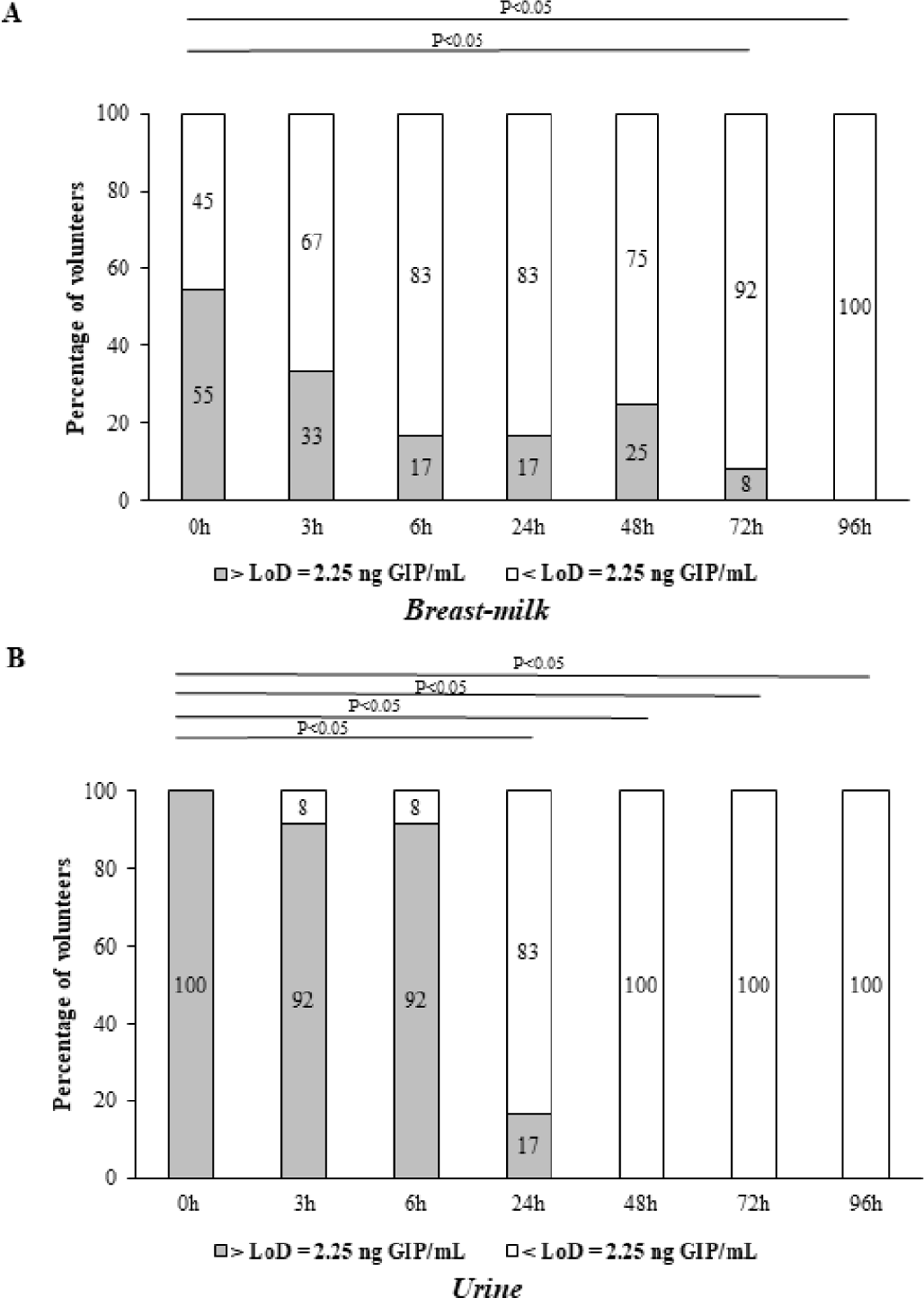
Comparison of GIP levels in breast milk and urine of 12 non-CD mothers on a GFD for over 96 h after gluten ingestion. (A) GIP secretion in breast milk; (B) GIP excretion in urine. Comparative analysis via the McNemar test of GIP levels before and after consumption of gluten. GIP, gluten immunogenic peptides; LoD, limit of detection.

## Discussion

To our knowledge, this study represents the first instance of identifying immunogenic gluten peptides in breast milk samples. The presence of GIP in breast milk indicates that gluten is absorbed through the intestinal mucosa, enters the bloodstream, and is subsequently secreted by the mammary gland. The detection of measurable amounts of GIP in breast milk suggests that infants are exposed to gluten antigens before their introduction into the diet, which could potentially impact CD development and other gluten-related disorders.

Our findings reveal that the secretion of GIP in breast milk exhibits dynamic and fluctuating changes over time, likely influenced by various factors such as maternal diet, hormonal fluctuations, and the feeding patterns of the infant, among others. This dynamic nature underscores the inherent complexity involved in studying GIP in breast milk. Following a gluten-containing diet challenge and the collection of multiple breast milk samples (ensuring at least one positive sample from each participant), the percentage of GIP-positive participants was to 75%. The secretion kinetics extended from 0 to 72 hours after the gluten-containing diet challenge and the commencement of the GFD. It is important to note that a considerable degree of intra- and interindividual variation was observed in the secretion of GIP, as previously reported for other food antigens [39–42].

Chirdo et al. [28] detected undegraded gliadins in all milk samples analyzed from lactating mothers following a normal diet, using a polyclonal antibody. The concentrations of gliadins observed exceeded those reported in our study to immunogenic gluten peptides. These variations may be attributed to differences in the study populations, the identified epitopes, and the specificity of the antibodies. The polyclonal antibodies are produced from a mixture of different immune cells, and they may bind to unrelated proteins, causing cross-reactivity. This cross-reactivity can lead to false signals or interference in assays. In contrast, our study has the advantage of detecting GIP using G12 and A1 moAbs, known for their reactivity with tandem epitopes contained in the major immunogenic peptides associated with CD [36,43].

As expected, in the cohort of mothers with CD on a GFD, the majority of the breast milk samples (82.6%, 19/23) tested negative for GIP. Only four breast milk samples tested positive for GIP. Although the mothers with CD in our study followed a GFD, the possibility of dietary lapses cannot rule out. A GFD is difficult to maintain, and adult patients with CD consume unsafe amounts of gluten on average [44–46]. Therefore, these results may be attributed to possible transgressions in the GFD as no food intake control was implemented for mothers with CD.

Understanding how other proteins manifest in breast milk is fundamental it provides crucial insights into the composition of breast milk and its role in infant nutrition and development. In our study, no significant differences were detected in the total protein levels in breast milk between non-CD and CD mothers. Thus, our findings suggest that a GFD does not influence variations in the levels of this macronutrient in breast milk. These results agree with those of prior studies that have demonstrated a remarkable consistency in the macronutrient composition of breast milk across different populations despite potential variations in maternal nutritional status [47]. With respect to food antigens, we also proved the transference of bovine casein into breast milk that represents one of the most commonly secreted non-human proteins [21]. The levels of this protein in breast milk were in the ng/mL range, similar to those of GIP and other food antigens [48]. No significant statistical differences in bovine casein levels were observed between the non-CD and CD cohorts. This result was expected, given that cow milk proteins are not only present in dairy products and their derivatives, but are also widely incorporated into a myriad of manufactured items such as bread, cold cuts, sausages, frozen fish, sweeteners, preserves, and medicines. Additionally, numerous additives used in the food industry are derived from milk [49].

The transfer of proteins from the maternal intestinal tract to the mammary gland is not yet well understood [24,48,50–52]. Additionally, food proteins secreted into the breast milk appear at highly variable concentrations at different time points following exposure and in different forms, either as free proteins or as components of immune complexes [24]. This makes it impossible to precisely predict the concentration at which food antigens/allergens will appear in breast milk and their impact on the immune response of the breast-fed children. For example, ovalbumin was detected in 68% of breast milk samples following egg ingestion, while beta-lactoglobulin was found in 62.5% of lactating women after consuming cow milk, exhibiting variable appearance and disappearance over a 15-hour sampling period. Another study observed the presence of beta-lactoglobulin in breast milk up to 7 days after oral intake of cow’s milk [37]. Kinetic analysis of peanut protein transfer into breast milk revealed that 11 out of 23 mothers excreted peanut proteins, with 73% showing appearance within 1 hour of ingestion. In this work, in a cohort of non-CD mothers, taking a single sample of breast milk and urine consecutively over time, we have demonstrated that the percentage of GIP-positive milk samples was significantly lower as compared to urine samples. The detection of GIP in the urine of 90% of non-CD mothers implied that the vast majority of volunteers in this cohort consumed gluten; however, this gluten intake was not reflected in breast milk (with only 24% breast milk being GIP positive). To understand this differential excretion/secretion GIP pattern between urine and breast milk and the excretion kinetics of gluten peptides in breast milk we designed a two-phase longitudinal trial: phase 1 involved a gluten-rich diet by menu planning, and phase 2 enforced a strict GFD. We measured GIP levels in the breast milk at various time points, ranging from 0 to 96 h after gluten ingestion and at the commencement of a GFD. The period of GIP excretion in urine after initiating the GFD was confined to the initial 24 h, with 100% of the volunteers testing positive for GIP during this period. In contrast, the period of GIP secretion in breast milk was extended to 72 h, with 75% of volunteers showing positive GIP results. This result confirmed the differential kinetics of GIP in breast milk and urine samples collected sequentially.

Gluten proteins are initially digested by the enzymes in the mother’s intestinal tract. However, this digestion is often incomplete, leaving behind resistant peptides that can cross the intestinal barrier. It is probable that these peptides reach the mammary glands throughout the entire lactation process. We hypothesize that these peptides may undergo partial degradation before being secreted into the mammary glands, potentially extending the time of secretion. Consequently, the mammary gland could serve as a reservoir for certain proteins, including gluten. This speculation arises from observations that mothers who followed the same diet, with menu planning, exhibited fluctuating gluten secretion and interindividual differences.

In this longitudinal trial, the children were breastfed during the milk sample collection period, and this may limit the scope of our results. It should be noted that not all breast milk samples could be collected within the designated 0 to 96 h timeframe, and GIP could have been secreted in some uncollected milk samples. Pooled 24h milk samples are generally considered gold standards for collection; however, they are not possible if the mother wants to maintain normal breastfeeding behaviors [42]. Furthermore, it is likely that the variations in the rate and concentration at which gluten is secreted into breast milk can be attributed to the complexity and content of meals consumed before or after gluten intake within our study group, and this could also contribute to the variations observed.

Our findings provide a conclusive demonstration of the presence of GIP in breast milk. Notable aspects of this study include the analysis of breast milk samples and the sequential analysis of urine samples using a specific assay designed for detecting GIP, coupled with the collection of multiple samples over several days. We now have the molecular tool to further investigate whether and how GIP are processed and secreted into breast milk. This investigation has raised important questions about the role that these proteins may play in the development of the immune system of infants, particularly considering that exposure to gluten in children occurs before the introduction of complementary feeding, as was previously understood. Therefore, future studies are necessary to know whether GIP act as sensitizing or tolerogenic agents in the immune system of breastfed babies.

## Author Contributions

Conceptualization, Á.R.-C., V.S., and I.C.; data curation, Á.R.-C and V.S.; formal analysis, Á.R.-C and V.S; investigation, Á.R.-C., V.S., M.d.L.M., C.C.-R., C.S. and I.C.; methodology, Á.R.-C., V.S., and I.C; resources, Á.R.-C., V.S., and I.C; writing—original draft, Á.R.-C., V.S., C.S. and I.C; writing—review and editing, Á.R.-C., V.S., M.d.L.M., C.C.-R., C.S. and I.C. All authors have read and agreed to the published version of the manuscript.

## Data Availability

All data produced in the present study are available upon reasonable request to the authors

## Acknowledgments

We would like to thank the Federation of Celiac Associations of Spain (FACE) and the Provincial Association of Celiacs of Seville (Asprocese) for their outreach to the population, as well as the volunteer mothers who enrolled in the study.

